# Differentiated service delivery models for HIV treatment in Malawi, South Africa, and Zambia: A landscape analysis

**DOI:** 10.1101/2020.08.25.20181818

**Authors:** Amy Huber, Sophie Pascoe, Brooke Nichols, Lawrence Long, Salome Kuchukhidze, Bevis Phiri, Timothy Tchereni, Sydney Rosen

## Abstract

**Introduction:** Many countries in Africa are scaling up differentiated service delivery (DSD) models for HIV treatment, but most existing data systems do not describe the models in use. We surveyed organizations that were supporting DSD models in 2019 in Malawi, South Africa, and Zambia to describe the diversity of DSD models being implemented at that time.

**Methods:** We interviewed DSD model implementing organizations for descriptive information about each of the organization’s models of care. We described the key characteristics of each model, including population of patients served, location of service delivery, frequency of interactions with patients, duration of dispensing, and cadre(s) of provider involved. To facilitate analysis, we refer to one organization supporting one model of care as an “organization-model.”.

**Results:** The 34 respondents (8 in Malawi, 16 in South Africa, 10 in Zambia) interviewed described a total of 110 organization-models, which included 19 facility based individual models, 21 out-of-facility based individual models, 14 healthcare worker led groups, and 3 client led groups; jointly, these encompassed 12 service delivery strategies. Over 2/3 (n=78) of the organization-models were limited to clinically stable patients. Almost all organization-models (n=96) continued to provide clinical care at established healthcare facilities; medication pickup took place at facilities, external pickup points, and adherence clubs. Required numbers of provider interactions per year varied widely, from a low of 2 to a high of 12. Dispensing intervals were typically 3 or 6 months in Malawi and Zambia and 2 months in South Africa. Individual models relied more on clinical staff (doctors, nurses, pharmacists), while group models made greater use of lay personnel (community health workers, counselors).

**Conclusions:** As of 2019, there was a large variety of differentiated service models being offered for HIV treatment in Malawi, South Africa, and Zambia, serving diverse patient populations.

## Introduction

In 2019, approximately 17.8 million people were receiving antiretroviral therapy (ART) for HIV in sub-Saharan Africa^1^. Achieving global targets for HIV treatment, known as “90-90-90^2^,” would require another 3 million patients to be added to the national HIV treatment programs. Meanwhile, donor spending in low and middle-income countries is declining, which has led countries, implementers, and funders to seek avenues of greater efficiency in service delivery^3,4^.

One response to this challenge is the development of “differentiated service delivery models” (DSD models) for HIV treatment^5^. DSD models, which typically reduce clinic visits and/or move services out of the clinic and may also alter the provider cadre and package of services provided, have multiple aims. They seek to make treatment more patient-centric by lessening the burden of frequent clinic visits; reduce costs to both the healthcare system and to patients; and sustain or improve clinical treatment outcomes^6^. DSD models are intended as an alternative to traditional or conventional care, which has typically been delivered at fixed-site clinics and requires at least four patient visits to the clinic per year, if not more. DSD models were originally targeted to adults in the general population who are “stable” on treatment, who comprise the largest population of patients, but models for other populations such as children and those with detectable viral loads have also been developed.

Although scale-up of DSD models is well underway in many African countries, existing data systems have not yet caught up with the diversity of approaches to HIV treatment delivery. Documentation of care delivered through DSD models is either not captured or is poorly captured in existing electronic medical record systems, and even paper-based patient files and site-level registers may not record, for example, whether a patient received a medication refill at the site, at a community pick-up point, or at home^7,8^. In most countries, moreover, a wider range of different models are being implemented than may be reflected in national HIV treatment guidelines, with both ministries of health and non-governmental partners designing and piloting approaches that vary from those in the guidelines. Some of these models are described in the published^9–12^ or unpublished^13,14^ literature or in funder or government databases, but most countries lack a comprehensive inventory of what is being tried.

In 2019, we undertook a series of interviews with DSD model implementing organizations in Malawi, South Africa, and Zambia to describe the current landscape of DSD model implementation in three high HIV prevalence countries in southern Africa, each representing a different income level (lower, upper-middle, and middle income, respectively). Interviews were conducted with as many non-governmental and governmental implementing organizations and agencies as could be identified. Here we present the information obtained through these interviews on the types and characteristics of DSD models underway. Our goal is to illustrate the diversity of models in use, identifying similarities and differences.

## Methods

We conducted cross-sectional structured interviews with DSD model implementing organizations in Malawi, South Africa, and Zambia. Each interview elicited the title and implementation start date for each differentiated model of care implemented by the organization and then collected descriptive information about the model, including population eligible, location and frequency of service delivery, provider cadre, and scale. Respondents were also asked for information on data and documentation, existing evaluations, and future plans for DSD projects.

### Identification of respondents

Our goal was to survey all the organizations that were either implementing or supporting implementation of DSD models in the study countries at the time of the survey (2019), whether for purposes of routine care, demonstration or pilot projects, or research studies. Potential survey respondents included government health agencies, implementing partners of national governments and of the U.S. President’s Emergency Plan for AIDS Relief (PEPFAR) and other donors, and other nongovernmental organizations. We first compiled a comprehensive list of potential respondents. We started with the study team’s own knowledge of each country’s HIV program and then supplemented this with recommendations from government DSD technical working groups and funding organization representatives. We then reviewed each country’s list with relevant national government representatives. Once the inventory of potential respondents was complete, each organization was invited to participate in the survey. Over the course of the survey, we also asked respondents to review and add to the list of potential participants from their own countries.

### Data collection

Data were collected using a semi-structured questionnaire administered by the study team in a face-to-face or electronic meeting with a representative of each participating organization. Interviews were audio-recorded and data entered into the project database after each interview was completed. Questions were limited to factual information about the DSD models being implemented. The interview instrument is included as Supplementary File 1. After all data had been entered, a report of the responses was sent to each respondent for verification, correction of errors, and provision of specific information that was not available at the time of the interview, such as the precise numbers of patients participating in each model.

### Data analysis

As this was a descriptive analysis, we aimed to group models within each country by their major characteristics, so that an overall profile of the models in use in each country could be developed. We first categorized the models using the taxonomy proposed by Grimsrud and colleagues^15^ and widely utilized within the DSD model literature. This taxonomy sorts models of care into four categories: facility based individual models (FBIM), such as fast-track clinic visits; out-of-facility based individual models (OFBIM), such as decentralized medication delivery; healthcare worker led groups (HCWLG), such as adherence clubs; and client led groups, such as community adherence groups or CAGs.

Although the four-category taxonomy described above is widely used, it also results in combining very diverse models, such as home delivery of ARVs and community-based clinical care, into the same category. We therefore also created a set of “strategies” that further refines the categories, grouping more similar models together. As we defined these strategies empirically, based on the interview responses, we regard them as results of the survey and present them in the results section.

We then used interview data to describe the key characteristics of each model, adapted from the well-known domains proposed by Duncombe et al^5^: population of patients served, location of service delivery, frequency of interactions with patients, duration of dispensing, and cadre(s) of provider involved (Supplementary Table 1). For each characteristic in the results section below, we start by describing traditional or conventional care, to provide a comparison with DSD model characteristics. We then report frequencies of each characteristic in each domain.

In reporting aggregate results, it is important to note that each organization-model combination was counted once, regardless of how many clinics offered the model, how many patients were enrolled, or whether other partners described the same model. For example, in this analysis a model being piloted at two clinics, with just a few dozen patients enrolled, and a model that had already been scaled up nationally and covered hundreds of thousands of patients were counted equally. Similarly, if two respondent organizations each responded that they were implementing the same model, this model was counted twice. Below, we refer to each combination of one model described by one respondent as an “organization-model.”

Although we did ask survey respondents to report the numbers of sites or facilities implementing each model and of patients participating, in most cases we were unable to obtain complete or accurate data for these. Where such numbers were available, it was generally not possible to confirm that no patients were double-counted in other implementer reports or that no individual patients were enrolled in more than one model. We therefore do not include information here on the scale of model implementation or coverage.

## Results

### Interviews conducted and models reported by respondents

We identified 36 potential respondents in the three countries and completed interviews with 34 of them. The remaining two, both in South Africa, declined to participate in the survey. Interviews were conducted between March 2019 and March 2020, with data verified by respondents between November 2019 and March 2020. We surveyed a total of 8 organizations in Malawi, 16 in South Africa, and 10 in Zambia.

The 34 respondents interviewed reported on a total of 110 organization-models, where an organization-model represents one organization supporting one model, as shown in the upper half of Table 1. Some models are specified in each country’s HIV treatment guidelines, to be scaled up nationally; others are bespoke models originating from nongovernmental organizations. Countries differed in their most commonly reported category of DSD model: more organizations in Malawi described facility-based individual models and more in Zambia described out-of-facility based individual models, while no organizations at all in South Africa reported supporting client-led group models.

**Table 1.** Numbers of organization-models described by respondents, by country, category, and strategy.

| Model category | Malawi | South Africa | Zambia | Total |
| --- | --- | --- | --- | --- |
| Total number of organization-models described | 26 | 43 | 41 | 110 |
| **Number of organization-models per country and category** |  |  |  |  |
| Facility based individual model | 13 | 6 | 13 | 32 |
| Out of facility based individual model | 5 | 22 | 16 | 43 |
| Health care worker led group | 7 | 15 | 4 | 26 |
| Client led group | 1 | 0 | 8 | 9 |
| **Numbers of organization-models per country and strategy** |  |  |  |  |
| Adherence clubs | 0 | 11 | 3 | 14 |
| Community adherence groups (CAG) | 1 | 0 | 8 | 7 |
| Community outreach | 3 | 3 | 4 | 11 |
| External pickup points | 2 | 17 | 6 | 26 |
| Extra clinic hours | 1 | 1 | 2 | 4 |
| Family models | 2 | 1 | 1 | 4 |
| Fast track services | 2 | 1 | 5 | 8 |
| Home delivery | 0 | 1 | 3 | 4 |
| Multimonth dispensing | 3 | 1 | 3 | 7 |
| Nonstable patient models | 6 | 4 | 1 | 11 |
| Key population models | 1 | 1 | 2 | 4 |
| Youth models | 5 | 2 | 3 | 10 |

We also grouped the 110 organization-models into twelve strategies, as described in Box 1 and listed alphabetically in the lower half of Table 1. The twelve strategies do not map directly onto the four categories; each taxonomy provides different information.

#### Box 1. DSD model strategies

| **Box 1. DSD model strategies** |  |
| --- | --- |
| *Adherence clubs* | Any group model that is led by a healthcare worker (professional or lay) |
| *Community adherence groups* | Any group model in which one patient picks up medications for other members of the group (typically abbreviated CAG) |
| *Community outreach* | A variety of models that bring both clinical care and medications into the community, such as nurse-led outreach |
| *External pickup points* | Any model that delivers ARVs to pickup points outside clinic facilities, such as lockers, community pharmacies, CCMDD decentralized pickup points, etc. |
| *Extra clinic hours* | Any model that adds additional hours to a facility's operations to facilitate access, such as on evenings or weekends |
| *Family models* | Any model designed to serve multiple and/or specific members of a family at once, such as a pediatric clinic or family clinic |
| *Fast track services* | Any model that creates a separate queue, kiosk, or procedure at a facility to speed up service delivery for stable patients |
| *Home delivery* | Any model that delivers ARVs to patients' homes, e.g. by a CHW or a bicycle courier |
| *Multimonth dispensing* | Any model in which the primary goal is to dispense medications for a longer duration than is done under standard care (usually 6 months) |
| *Nonstable patient models* | Models for patients who do not meet definitions of clinical stability, such as high viral load clinics and advanced disease clinics |
| *Key population models* | Models for a key population such as MSM or FSW |
| *Youth models* | Any model specifically for youth/teens/adolescents, e.g. teen clubs in Malawi and the scholar model in Zambia |

There are several provisos to model taxonomies in Table 1. First, while we used both the names given to the models by the implementing organizations and interview respondents’ descriptions of the models to allocate each organization-model to a category and approch, in some cases we were uncertain and had to go with what appeared to be the closest fit, given what we knew. Second, not all of the models listed in Table 1 are mutually exclusive. Six-month dispensing, for example, can be implemented within many other models of care. Third, many models also provide some services that are not strictly consistent with their model category or approach. Out of facility based individual models, for example, may provide some services at facilities, while facility-based models may include home visits for patients who miss appointments; both group model categories likely include some individual services. Similarly, community outreach strategies may incorporate external medication pickup points, along with community-based clinical care. Finally, a model that is considered “differentiated” in one country—like three-month dispensing in South Africa—and is thus included in Table 1 may be regarded as standard of care in another, like Malawi, for which it is not mentioned in Table 1.

### Populations served

As anticipated, most of the models described in the survey served adults in the general population who were stable on treatment. Definitions of stability varied by country and model, but most included a minimum of 12 months on ART and evidence of viral suppression. A number of other models were designed for people with advanced HIV disease, an unsuppressed viral load, or newly initiated on ART. Models also existed for different age groups (children, adolescents) and vulnerability groups (men who have sex with men (MSM), female sex workers (FSW), pregnant women). We note that the survey was completed prior to the adoption of COVID-19 related changes to eligibility criteria, which may or may not be permanent. In Table 2, the number of organization-models for each population group is reported by country.

**Table 2.**
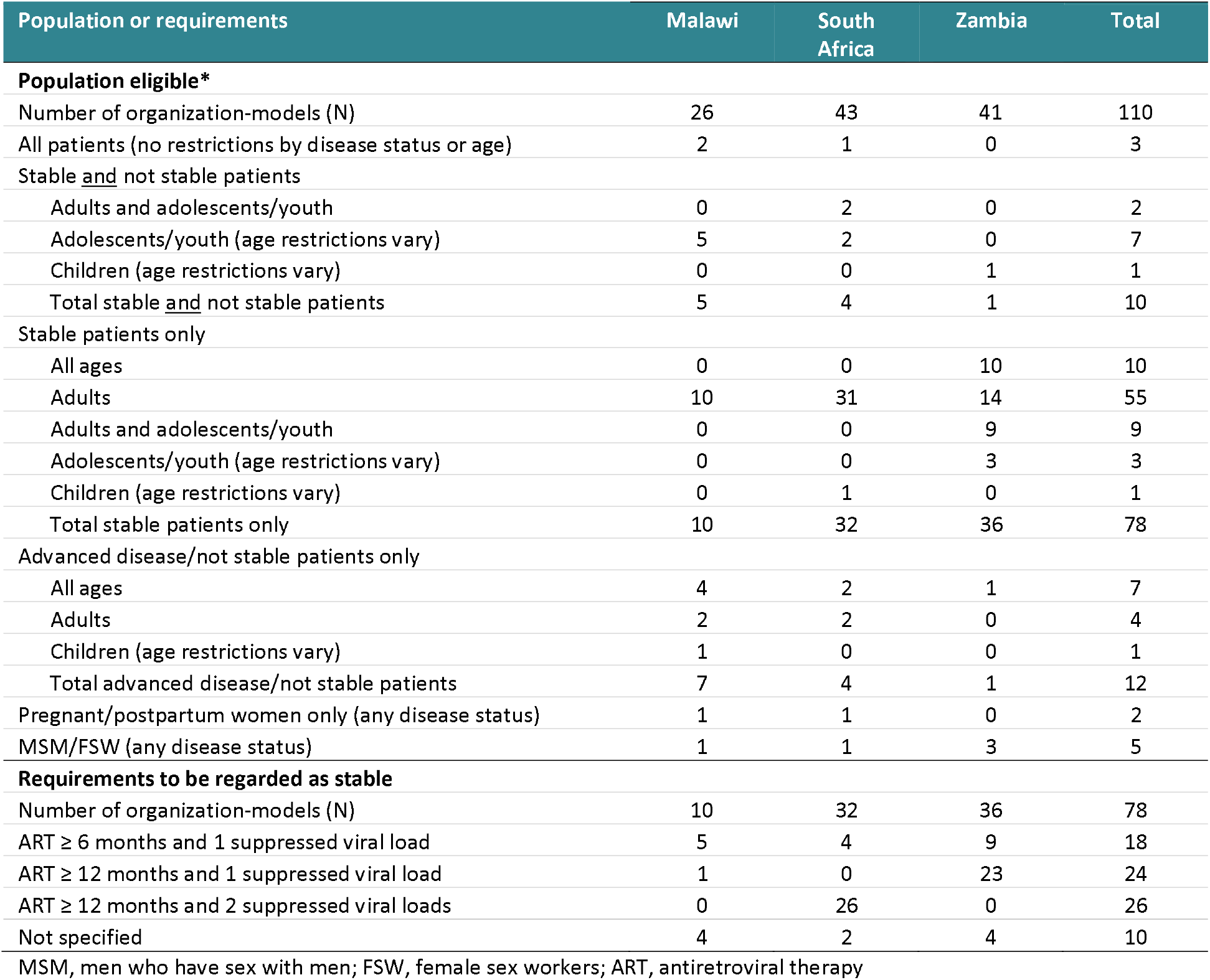
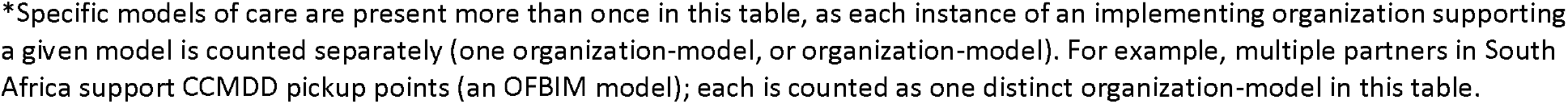
Populations served by current DSD models and criteria for defining stability, by country.

| Population or requirements | Malawi | South Africa | Zambia | Total |
| --- | --- | --- | --- | --- |
| **Population eligible\*** |  |  |  |  |
| Number of organization-models (N) | 26 | 43 | 41 | 110 |
| All patients (no restrictions by disease status or age) | 2 | 1 | 0 | 3 |
| Stable <u>and</u> not stable patients |  |  |  |  |
| Adults and adolescents/youth | 0 | 2 | 0 | 2 |
| Adolescents/youth (age restrictions vary) | 5 | 2 | 0 | 7 |
| Children (age restrictions vary) | 0 | 0 | 1 | 1 |
| Total stable <u>and</u> not stable patients | 5 | 4 | 1 | 10 |
| Stable patients only |  |  |  |  |
| All ages | 0 | 0 | 10 | 10 |
| Adults | 10 | 31 | 14 | 55 |
| Adults and adolescents/youth | 0 | 0 | 9 | 9 |
| Adolescents/youth (age restrictions vary) | 0 | 0 | 3 | 3 |
| Children (age restrictions vary) | 0 | 1 | 0 | 1 |
| Total stable patients only | 10 | 32 | 36 | 78 |
| Advanced disease/not stable patients only |  |  |  |  |
| All ages | 4 | 2 | 1 | 7 |
| Adults | 2 | 2 | 0 | 4 |
| Children (age restrictions vary) | 1 | 0 | 0 | 1 |
| Total advanced disease/not stable patients | 7 | 4 | 1 | 12 |
| Pregnant/postpartum women only (any disease status) | 1 | 1 | 0 | 2 |
| MSM/FSW (any disease status) | 1 | 1 | 3 | 5 |
| **Requirements to be regarded as stable** |  |  |  |  |
| Number of organization-models (N) | 10 | 32 | 36 | 78 |
| ART ≥ 6 months and 1 suppressed viral load | 5 | 4 | 9 | 18 |
| ART ≥ 12 months and 1 suppressed viral load | 1 | 0 | 23 | 24 |
| ART ≥ 12 months and 2 suppressed viral loads | 0 | 26 | 0 | 26 |
| Not specified | 4 | 2 | 4 | 10 |

MSM, men who have sex with men; FSW, female sex workers; ART, antiretroviral therapy
\*Specific models of care are present more than once in this table, as each instance of an implementing organization supporting a given model is counted separately (one organization-model, or organization-model). For example, multiple partners in South Africa support CCMDD pickup points (an OFBIM model); each is counted as one distinct organization-model in this table.

Half (n=55, 50%) of the organization-models were limited to stable adults, and over two thirds (n=78, 71%) to stable patients overall. About a quarter (n=25, 23%) either accepted advanced disease or unsuppressed patients along with stable patients or were explicitly designed for those with advanced disease or viral failure. A small handful targeted special populations (n=7, 6%).

As mentioned above, where “stability” was a criterion for DSD model enrollment, definitions varied. In Table 2, we also summarize the criteria used for the 78 organization-models limited to stable patients. Of the 70 organization-models for which stability criteria were specified, more than two thirds (n=50, 71%) required that patients have spent at least a full year on ART prior to DSD model eligibility, and all required at least one suppressed viral load measurement.

### Location

Locations of models reported by survey respondents are shown in Table 3. As mentioned above, one of the main criteria for differentiating HIV treatment delivery from traditional, clinic-based care is the location of service. Prior to differentiation, nearly all care and medication dispensing took place at fixed-site clinics, with occasional community outreach efforts to trace defaulters or provide treatment education or adherence support. DSD models offer services in a wide range of locations, from fixed-site clinics to private pharmacies and community meeting spaces, to patients’ homes. For models that provide most services off site, patients typically remain the responsibility of a fixed-site clinic, which supervises the delivery of care through the alternative model and maintains patient records.

**Table 3.** Number of organization-models by location of service delivery and country.

| Location of service delivery | Number of organization-models |  |  |  |
| --- | --- | --- | --- | --- |
|  | Malawi<br>N=26 | South<br>Africa<br>N=43 | Zambia<br>N=41 | Total<br>N=110 |
| Facility for clinical care with medication pickup at: |  |  |  |  |
| Facility | 19 | 6 | 16 | 41 |
| External pick-up point | 4 | 18 | 15 | 37 |
| Adherence club | 0 | 13 | 1 | 14 |
| Home | 0 | 1 | 3 | 4 |
| Total (facility location for clinical care) | 23 | 38 | 35 | 96 |
| External location for clinical care: |  |  |  |  |
| Clinical care and medications in community | 3 | 3 | 5 | 11 |
| Clinical care and medications at adherence club | 0 | 2 | 0 | 2 |
| Clinical care and medications at home | 0 | 0 | 1 | 1 |
| Total (external location for clinical care) | 3 | 5 | 6 | 14 |

Almost all of the organization-models (n=96, 87%) continued to provide clinical care at established healthcare facilities, though each country has a few organization-models that delivered clinical care outside the facility. Medication pickup locations vary by country. Facility-based pickup was most common among organization-models in Malawi; external pickup points and pickup at adherence clubs, frequently located at the facility rather than in the community, were widely utilized in South Africa; and medication pickup at facilities and at external pickup points were both common in Zambia.

### Frequency of interactions with healthcare system

In addition to location of service delivery, the number of times per year that a patient must interact with the healthcare system—either an established clinic or an off-site location—is a critical differentiator of the alternative models of care that have been developed. During the first decade of large-scale, public sector ART programs in Africa, patients typically collected their medications once a month, with clinical monitoring conducted 4-12 times per year. Dispensing intervals expanded gradually, from a maximum of one month at a time to up to three months in some countries, but frequent clinic visits remained the norm.

In an effort to reduce the burden of treatment on patients and clinics, where time and space should be freed up by requiring fewer ART visits per year, DSD models generally try to keep stable patients out of the clinic. This may be done by reducing the absolute number of clinic visits per year and/or replacing some or all traditional or “full” clinic visits with briefer visits for medication pickup only, such as fast-track visits, or off-site interactions, such as adherence club participation or medication access at an external pickup point. Table 4 summarizes the number of clinic and DSD model interactions required per year, with “clinic visit” referring to a full or traditional visit and “DSD interaction” including short clinic visits or medication refill/pick-up visits that were designed for a DSD model. Models that required evidence of viral suppression for eligibility are reported separately from those that did not.

**Table 4.**
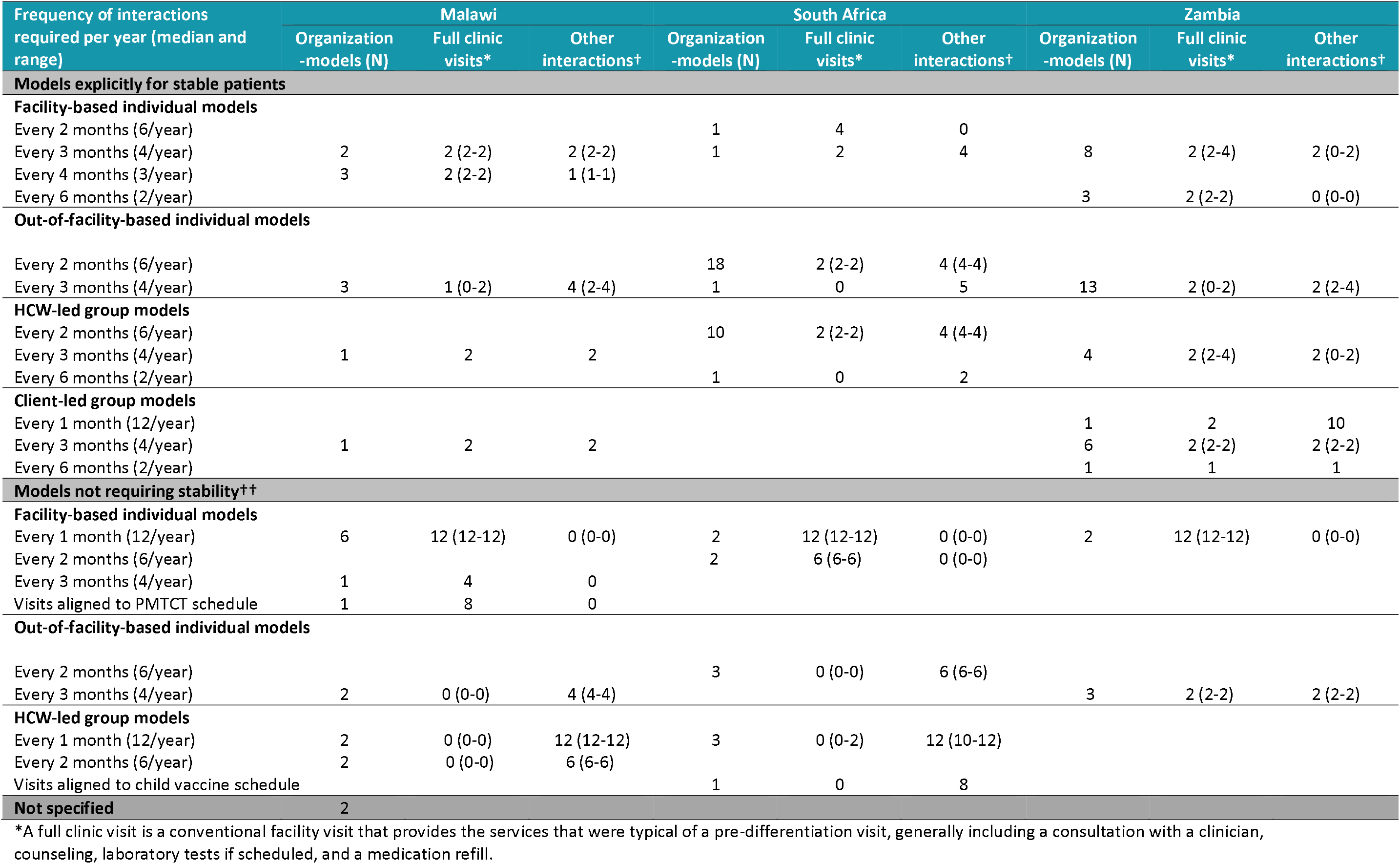
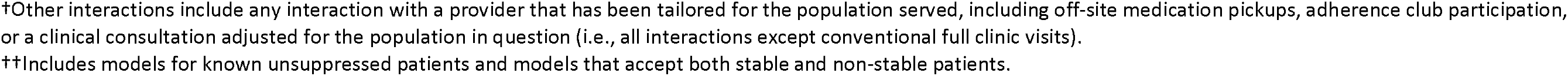
Frequency of healthcare system interactions per year, by country, model category, and viral suppression eligibility criterion.

| Frequency of interactions required per year (median and range) | Malawi |  |  | South Africa |  |  | Zambia |  |  |
| --- | --- | --- | --- | --- | --- | --- | --- | --- | --- |
|  | Organization -models (N) | Full clinic visits* | Other interactions† | Organization -models (N) | Full clinic visits* | Other interactions† | Organization -models (N) | Full clinic visits* | Other interactions† |
| **Models explicitly for stable patients** |  |  |  |  |  |  |  |  |  |
| **Facility-based individual models** |  |  |  |  |  |  |  |  |  |
| Every 2 months (6/year) |  |  |  | 1 | 4 | 0 |  |  |  |
| Every 3 months (4/year) | 2 | 2 (2-2) | 2 (2-2) | 1 | 2 | 4 | 8 | 2 (2-4) | 2 (0-2) |
| Every 4 months (3/year) | 3 | 2 (2-2) | 1 (1-1) |  |  |  |  |  |  |
| Every 6 months (2/year) |  |  |  |  |  |  | 3 | 2 (2-2) | 0 (0-0) |
| **Out-of-facility-based individual models** |  |  |  |  |  |  |  |  |  |
| Every 2 months (6/year) |  |  |  | 18 | 2 (2-2) | 4 (4-4) |  |  |  |
| Every 3 months (4/year) | 3 | 1 (0-2) | 4 (2-4) | 1 | 0 | 5 | 13 | 2 (0-2) | 2 (2-4) |
| **HCW-led group models** |  |  |  |  |  |  |  |  |  |
| Every 2 months (6/year) |  |  |  | 10 | 2 (2-2) | 4 (4-4) |  |  |  |
| Every 3 months (4/year) | 1 | 2 | 2 |  |  |  | 4 | 2 (2-4) | 2 (0-2) |
| Every 6 months (2/year) |  |  |  | 1 | 0 | 2 |  |  |  |
| **Client-led group models** |  |  |  |  |  |  |  |  |  |
| Every 1 month (12/year) |  |  |  |  |  |  | 1 | 2 | 10 |
| Every 3 months (4/year) | 1 | 2 | 2 |  |  |  | 6 | 2 (2-2) | 2 (2-2) |
| Every 6 months (2/year) |  |  |  |  |  |  | 1 | 1 | 1 |
| **Models not requiring stability††** |  |  |  |  |  |  |  |  |  |
| **Facility-based individual models** |  |  |  |  |  |  |  |  |  |
| Every 1 month (12/year) | 6 | 12 (12-12) | 0 (0-0) | 2 | 12 (12-12) | 0 (0-0) | 2 | 12 (12-12) | 0 (0-0) |
| Every 2 months (6/year) |  |  |  | 2 | 6 (6-6) | 0 (0-0) |  |  |  |
| Every 3 months (4/year) | 1 | 4 | 0 |  |  |  |  |  |  |
| Visits aligned to PMTCT schedule | 1 | 8 | 0 |  |  |  |  |  |  |
| **Out-of-facility-based individual models** |  |  |  |  |  |  |  |  |  |
| Every 2 months (6/year) |  |  |  | 3 | 0 (0-0) | 6 (6-6) |  |  |  |
| Every 3 months (4/year) | 2 | 0 (0-0) | 4 (4-4) |  |  |  | 3 | 2 (2-2) | 2 (2-2) |
| **HCW-led group models** |  |  |  |  |  |  |  |  |  |
| Every 1 month (12/year) | 2 | 0 (0-0) | 12 (12-12) | 3 | 0 (0-2) | 12 (10-12) |  |  |  |
| Every 2 months (6/year) | 2 | 0 (0-0) | 6 (6-6) |  |  |  |  |  |  |
| Visits aligned to child vaccine schedule |  |  |  | 1 | 0 | 8 |  |  |  |
| **Not specified** | 2 |  |  |  |  |  |  |  |  |

\*A full clinic visit is a conventional facility visit that provides the services that were typical of a pre-differentiation visit, generally including a consultation with a clinician, counseling, laboratory tests if scheduled, and a medication refill.
†Other interactions include any interaction with a provider that has been tailored for the population served, including off-site medication pickups, adherence club participation, or a clinical consultation adjusted for the population in question (i.e., all interactions except conventional full clinic visits).
††Includes models for known unsuppressed patients and models that accept both stable and non-stable patients.

While the total number of clinic visits and DSD interactions required per year varied widely by model, most organization-models continued to expect patients to interact with healthcare providers, either at or outside the facility, at least 4 times per year.

### Dispensing intervals

Interaction frequency, as presented in Table 4, appears to be determined in part, but not solely, by duration of dispensing; if patients receive a three-month supply of ARV medications at a time, interactions must take place at least quarterly. Many models are designed to interact with patients more frequently than once per quarter, however. As mentioned above, early in national ART programs, patients received a maximum of one month of medications at a time, and refills were dispensed only by fixed-site clinics. Since a medication refill visit, even if a clinical consultation is not required, often takes a full day due to long waiting times, a promising way to improve treatment delivery is to dispense more months at a time. “Multi-month dispensing” (i.e., at least three months of medication dispensed) is now the norm in Zambia and Malawi and under consideration in South Africa, but the number of months allowed in our survey varied from two to six. Table 5 summarizes dispensing intervals expected for DSD models in each country.

**Table 5.** Months of ART dispensed.

| Average months of ARVs dispensed in model | Number of organization-models |  |  |  |
| --- | --- | --- | --- | --- |
|  | Malawi | South Africa | Zambia | Total |
| 1 month | 7 | 5 | 3 | 15 |
| 2 months | 2 | 34 | 0 | 36 |
| 3 months | 7 | 1 | 12 | 20 |
| 1 or 3 months (patients typically start with 1 month, then move to 3 months) | 2 | 2 | 2 | 6 |
| 6 months | 3 | 1 | 7 | 11 |
| 3 or 6 months (typically previously dispensed 3 months but transitioning to 6 months in line with national policy) | 3 | 0 | 17 | 20 |
| Not reported | 2 | 0 | 0 | 2 |

Dispensing intervals varied in Malawi, with some organization-models dispensing only one month at a time and others offering six months per pickup. This variation in dispensing is related to patient population served, with special populations and those with an unsuppressed viral load generally receiving shorter intervals and stable patients generally receiving 3 or 6 months. In South Africa, two-month dispensing remained the norm, with only one six-month dispensing model reported. Dispensing intervals in Zambia reflected the transition underway at the time of the survey between a standard of care of 3 months/pickup to 6 month dispensing, which is now national policy for stable patients.

### Providers

The final characteristic that helps to describe DSD models is the cadre of staff that provides services. Task-shifting from more to less senior clinical staff, and from clinical staff to lay providers, has been common for many years. Some of the DSD models described by survey respondents developed this practice further, relying more heavily on non-clinical providers located outside facilities, while others continued to make use of different cadres of clinical providers. Table 6 describes the cadres providing clinical care and ARV dispensing in each category of model.

**Table 6.** Clinical care provider and ART dispenser.

| Provider (number of organization-models) | Facility based individual (FBIM) | Out of facility individual (OFBIM) | Healthcare worker led group (HCWLG) | Client led group (CLG) | Total |
| --- | --- | --- | --- | --- | --- |
| **Clinical care provider** |  |  |  |  |  |
| Medical doctor/medical officer/clinical officer | 9 | 1 | 2 | 2 | 14 |
| Nurse | 7 | 28 | 16 | 0 | 51 |
| Community health worker | 0 | 1 | 0 | 1 | 2 |
| Non-specified clinician | 3 | 7 | 3 | 2 | 15 |
| Unclear/not reported | 13 | 6 | 5 | 4 | 28 |
| **ART dispenser** |  |  |  |  |  |
| Pharmacist/pharmacy assistant | 12 | 7 | 2 | 0 | 21 |
| Nurse/clinician | 7 | 7 | 3 | 0 | 17 |
| Community health worker | 0 | 6 | 5 | 0 | 11 |
| Designated patient | 0 | 2 | 1 | 9 | 12 |
| Lay counselor/trained non-clinical personnel | 0 | 14 | 9 | 0 | 23 |
| Medication locker/remote pharmacist | 0 | 3 | 0 | 0 | 3 |
| Unclear/not reported | 13 | 4 | 6 | 0 | 23 |

In general, individual models relied more on clinical staff (doctors, nurses, and pharmacists), while group models made greater use of lay personnel (community health workers and counselors). The cadre providing these services though was frequently not reported, however, particularly for facility-based individual models.

## Discussion

This survey of organizations implementing differentiated service delivery models for HIV treatment revealed 110 instances of DSD model provision in Malawi, South Africa, and Zambia in 2019. Three of the four commonly seen categories of DSD models—facility-based individual care, out-of-facility-based individual care, and healthcare worker led groups—were well represented in all three countries; client-led groups were common only in Zambia. The categories fell fairly naturally into 12 strategies for service delivery which, as a set, may offer a more specific way to describe DSD models for HIV treatment going forward. Even within these strategies, however, specific models did vary, particularly in terms of specific populations targeted and locations and timing of medication pickup or delivery. Most models continued to provide clinical care at facilities and, as anticipated, most models were limited to stable adult patients.

Although DSD models are often assumed to be “less intensive” approaches to service delivery, the models being implemented in 2019 still required relatively frequent interaction between patients and providers. Four interactions per year was most common for stable patients; models that allowed or focused on non-stable patients generally required more interactions. Dispensing intervals also varied by model and country. Models in Zambia and Malawi were beginning to reflect these countries’ adoption of six-month dispensing policies, a process that has likely been hastened by COVID-19, while dispensing intervals remained relatively short (2-3 months) in South Africa.

DSD models are also assumed to incorporate task-shifting from more- to less-trained cadres of service providers. In most of the models reported by survey respondents, though, formally trained clinical staff (doctors, nurses, pharmacists) continued to provide the majority of services, even in out-of-facility based models.

While we attempted to generate a comprehensive description of DSD models in use in 2019, our survey had many limitations. First, it is possible that our survey missed some implementing organizations in our target countries, and it is unlikely that the inventory of models in Table 3 is truly comprehensive. It includes all the DSD models mentioned by the partners interviewed, but there are almost certainly other approaches being tried by others. We are confident that models that are missing from Table 4, however, are relatively small and/or new initiatives at the time of the survey. A more important limitation is that the information collected pertains to the situation in 2019, when the survey was conducted. The world of differentiated service delivery is evolving rapidly. Some of the models described to us in 2019 almost certainly no longer exist one year later, while new models that had not yet been launched in 2019 may be underway now. Similarly, government guidelines for the models being rolled out nationally in each country may also have been updated since the survey was conducted.

Finally, we were not able to weight the models described to us by their importance within national DSD landscapes. As mentioned above, while we attempted to obtain estimates of numbers of facilities and patients participating in each model, results were incomplete and difficult to interpret. We thus could not reliably distinguish between a bespoke, respondent-specific model serving just a handful of patients and a well-established, national model serving tens of thousands. Our data captured the range of diversity, but not its scale.

Despite these limitations, the survey reported here provides what we believe is the most complete description available yet of DSD models for HIV treatment in southern Africa. It can both provide examples to other countries of new approaches they have not yet considered and serve as a baseline of model diversity, against which to evaluate the further development of differentiated service delivery in the coming years.

## Data Availability

The data set used in this manuscript will be anonymized and made available through a repository such as Dryad.com 12 months after the manuscript is published. Until then, data may be available on request from the corresponding author.

## Supplementary materials

**Supplementary table 1.** DSD domains

| Domain | Question asked | Definition |
| --- | --- | --- |
| Population | Who does the model serve? | Population is categorized by patient ART status (stable, newly initiated, not stable) and by age (child, adolescent, adult); occasionally also by vulnerability group (general, men who have sex with men (MSM), adolescents etc.). |
| Location | Is care provided in the clinic or off-site? | Location is categorized as 1) receiving all services at a fixed health care facility, 2) receiving all services outside of health care facility; or 3) mixed, receiving some services in each location. |
| Frequency | How often does the patient interact with a healthcare provider or facility? | Number of unique times a client receives any service per 12-month period. For example, a client receiving a combined clinical consultation and drug pickup at the same place on the same day every 3 months has a frequency of 4. |
| Dispensing interval | How many months of medications are dispensed? | Number of months of ARV medications provided at each interaction with the healthcare system. This indicator measures dispensing, not prescribing (e.g. a six-month prescription delivered twice, in three-month quantities each, has a three month duration). |
| Provider | Which cadres of clinical and/or lay staff provide the services? | Potential cadres include doctors, nurses, lay counselors, community health workers, pharmacists, and others designated within the models. |

Supplementary file 1. Survey instrument

## References

1. World Health Organization. Latest HIV estimates and updates on HIV policies uptake, July 2020. https://www.who.int/hiv/data/2019_summary-global-hiv-epi.png.

2. UNAIDS. 90-90-90: An ambitious treatment target to help end the AIDS epidemic. Published 2014. https://www.unaids.org/en/resources/documents/2017/90-90-90.

3. Barker C, Dutta A, Klein K. Can differentiated care models solve the crisis in HIV treatment financing? Analysis of prospects for 38 countries in sub-Saharan Africa. J Int AIDS Soc. 2017;20(5):68–79.

4. Kates J, Wexler A, Lief E. Financing the response to HIV in low- and middle-income countries: International assistance from donor governments in 2015. Published 2016. https://www.unaids.org/sites/default/files/media_asset/financing-the-response-to-HIV-in-low-and-middle-income-countries_en.pdf.

5. Duncombe C, Rosenblum S, Hellmann N, et al. Reframing HIV care: putting people at the centre of antiretroviral delivery. Trop Med Int Heal. 2015;20(4): 430–47.

6. Grimsrud A, Bygrave H, Doherty M, et al. Reimagining HIV service delivery⍰: the role of differentiated care from prevention to suppression. J Acquir Immune Defic Syndr. 2016;(19):10–12.

7. Ehrenkranz PD, Calleja JMG, El-sadr W, et al. A pragmatic approach to monitor and evaluate implementation and impact of differentiated ART delivery for global and national stakeholders. J Int AIDS Soc. 2018;21(3):e25080.

8. Reidy WJ, Rabkin M, Syowai M, Schaaf A, El-Sadr WM. Patient-level and program-level monitoring and evaluation of differentiated service delivery for HIV⍰: a pragmatic and parsimonious approach is needed. AIDS. 2018 32(3): 399–401.

9. Rabkin M, Strauss M, Mantell JE, et al. Optimizing differentiated treatment models for people living with HIV in urban Zimbabwe: Findings from a mixed methods study. PLoS One. 2020;15(1): e0228148.

10. Hanrahan CF, Schwartz SR, Mudavanhu M, et al. The impact of community-versus clinic-based adherence clubs on loss from care and viral suppression for antiretroviral therapy patients⍰: Findings from a pragmatic randomized controlled trial in South Africa. PLOS Med. 2019;16(5):e1002808.

11. Ehrenkranz P, Grimsrud A, Rabkin M. Differentiated service delivery: Navigating the path to scale. Curr Opin HIV AIDS. 2019;14(1):60–65.

12. Phiri K, Mcbride K, Siwale Z, et al. Provider experiences with three- and six-month antiretroviral therapy dispensing for stable clients in Zambia. AIDS Care. 2020; https://doi.org/10.1080/09540121.2020.1755010

13. Kuchukhidze S, Long L, Pascoe S, et al. Differentiated models of service delivery (DSD) for antiretroviral treatment of HIV in Sub-Saharan Africa: a review of the gray literature as of June 2019. https://sites.bu.edu/ambit/files/2019/11/AMBIT-report-03-gray-literature-review-2019-11-08.pdf.

14. Long L, Kuchukhidze S, Pascoe S, et al. Retention in care and viral suppression in differentiated service delivery models for HIV treatment in sub-Saharan Africa: a rapid systematic review. Preprints. 2020;(May):1–21.

15. Grimsrud A, Barnabas R V., Ehrenkranz P, Ford N. Evidence for scale up: The differentiated care research agenda. J Int AIDS Soc. 2017; 20(Suppl 4): 22024.

